# Online survey of treatment experiences for obsessive-compulsive disorder

**DOI:** 10.1101/2022.10.21.22281372

**Authors:** E.J. Kirkham, Y. Cao, M. Król

## Abstract

**Introduction:** There is a lack of knowledge about the treatment experiences of people living with obsessive-compulsive disorder (OCD).

**Method:** Participants with OCD (*n* = 202) took part in an online survey in which they answered questions about what treatment they had received. They also completed measures of current OCD, anxiety and depression.

**Results:** Scores above the clinical cut-off for OCD were high (> 70%) across the sample, irrespective of what kind of treatment had been received. Despite this, most participants felt their OCD was better now than it had been pre-treatment. Mean OCD scores were similar between treatment groups (OCD-focused, partly OCD-focused, non-OCD focused, no treatment), though there was some evidence that OCD-focused treatment was associated with lower OCD scores, especially for symptoms of hoarding, neutralising and ordering.

**Discussion:** These findings contribute to evidence of unmet need in OCD care. Practitioners should be aware that OCD is often a chronic condition which requires specialist care, and may require more than one course of treatment.

---

Obsessive-compulsive disorder (OCD) is often a chronic condition, with relatively high levels of treatment non-response and relapse (Del Casale et al., 2019; Visser et al., 2014). Despite this, research studies typically focus on whether a one specific course of treatment is effective, and usually include relatively short-term follow-ups of the participants. Subsequently there is a lack of knowledge about how people living with OCD, many of whom do not achieve remission (Pittenger & Bloch, 2014; Sookman et al., 2021), experience the treatments offered to them in the “real world”. This brief communication therefore seeks to provide an overview of the treatment experience of a group of people with OCD, and to detail the findings of exploratory analyses on whether prior treatment experiences are related to current OCD symptomatology.

## Method

The research received ethical approval from the Philosophy, Psychology and Language Sciences Research Ethics Committee of the University of Edinburgh (Reference Number: 227-2122/11). All participants provided informed consent prior to taking part. Recruitment was primarily conducted through social media and Prolific Academic (www.prolific.co). Those who took part through Prolific Academic received an average of £2.25, while a £4 per-participant donation to an OCD charity was made on behalf of the rest of the sample.

An online survey was used to collect data on variables associated with OCD. The survey contained a series of questions designed to provide pilot quantitative data on treatment experience, as well as pre-existing questionnaires including the Obsessive-Compulsive Inventory Revised (OCI-R; Foa et al., 2002) and the Hospital Anxiety and Depression Scale (HADS; Zigmond & Snaith, 1983).

The survey was completed by 390 participants, 202 of whom reported that they had OCD. The present report focuses on this sample of participants with OCD. Amongst this sample, 66.2% were female, 32.3% were male and 1.5% selected non-binary or “other”. The mean age was 29.9 years (*SD* = 10.0), and the majority (84.1%) of participants had attended university. Ethnicity was as follows: 8.5% Asian, 9.5% Black, 5.0% mixed or multiple ethnic groups, 72.6% White and 4.5% other ethnic group.

A one-way ANCOVA was used to examine the effect of treatment type on OCD score when controlling for anxiety and depression. Significant effects were followed up with pairwise comparisons. Following this, a mixed-model ANCOVA was used to examine whether scores on different OCD subscales varied according to whether treatment was fully focused on OCD or only partly focused on OCD. Significant effects were examined using pairwise comparisons and univariate ANCOVAs. Multiple linear regression was used to examine whether treatment response was associated with any of the six OCD subscales.

## Results

### OCD experience

Within the sample of people who stated that they had OCD, 90 had received a diagnosis and 112 had not. The mean OCI-R score was higher for those without a diagnosis (*M* = 36.68, *SD* = 13.73) than those with a diagnosis (*M* = 32.81, *SD* = 14.40), though this difference did not reach significance (*t*(197) = -1.94, *p* = .05). The proportion of individuals who scored above the clinical cut-off on the OCI-R (>= 21) was 78.4% for the diagnosed group and 82.9% for the non-diagnosed group. All participants (irrespective of whether or not they had a diagnosis) were asked to select one of four statements which best described the course of their OCD symptoms (Figure 1). The majority indicated that the severity of their symptoms varied over time.

**Figure 1:**
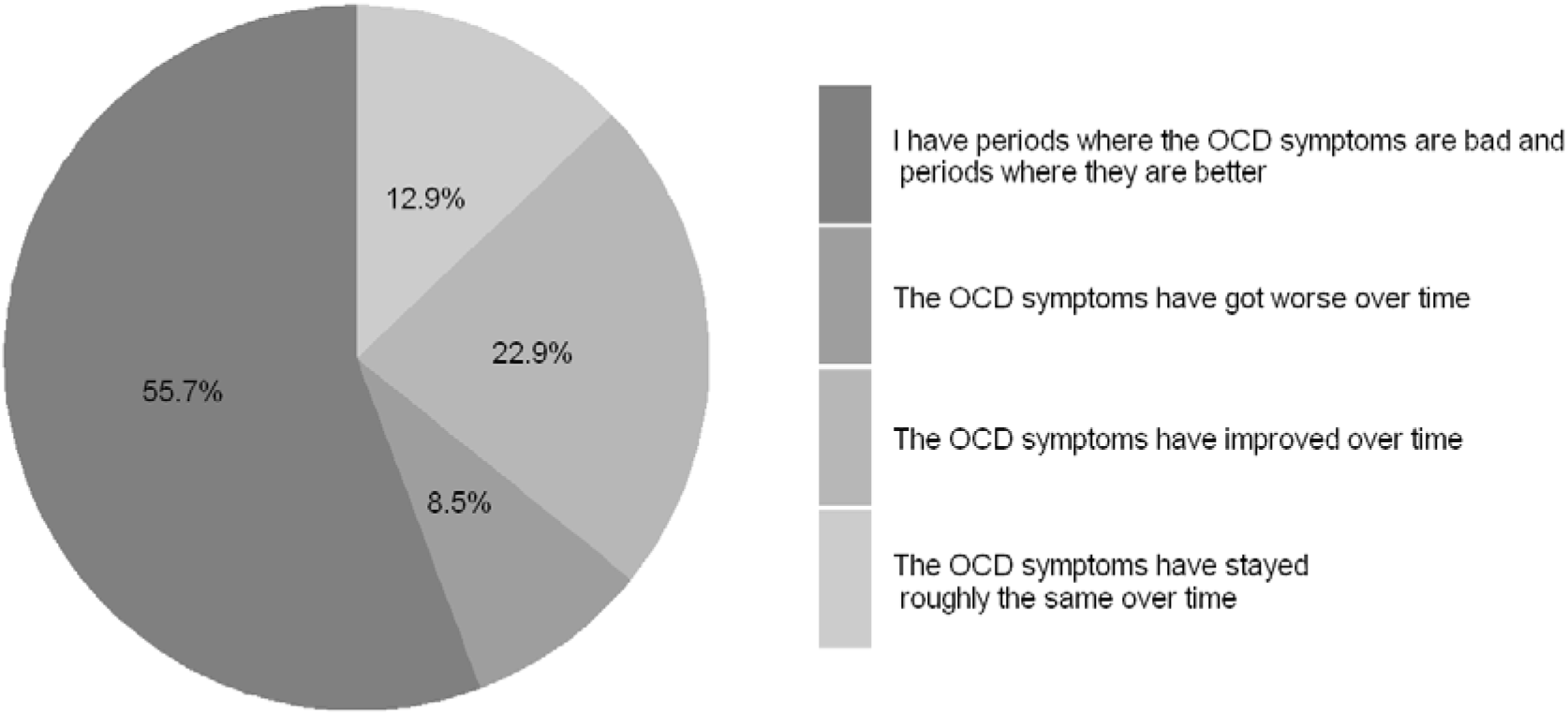
Experience of OCD symptoms.

### OCD symptoms by type of mental health treatment received

A one-way ANCOVA was used to examine the effect of treatment type on OCI-R score when controlling for anxiety and depression. Amongst people with self-reported OCD (diagnosed or not diagnosed; *n* = 186) there was a significant effect of treatment type on OCI-R score, Figure 2; *F*(5, 180) = 2.66, *p* = .02). Pairwise comparisons with Bonferroni correction indicated that people who had received OCD-specific treatment had significantly lower OCI-R scores than people who had received mental health treatment that did not address OCD at all (mean difference = -7.55, *p* = .05, 95% CI -15.09, -.001). No other comparisons between groups reached significance. Notably this means that those who had received mental health treatment, whether focused on OCD or not, had comparable OCD scores to those who had not received any mental health treatment. The percentage of people scoring above the clinical threshold for current OCD was 70.8% for OCD treatment, 84.9% for mental health treatment partly focused on OCD, 86.0% for mental health treatment with no OCD focus, and 74.3% for no mental health treatment.

**Figure 2.**
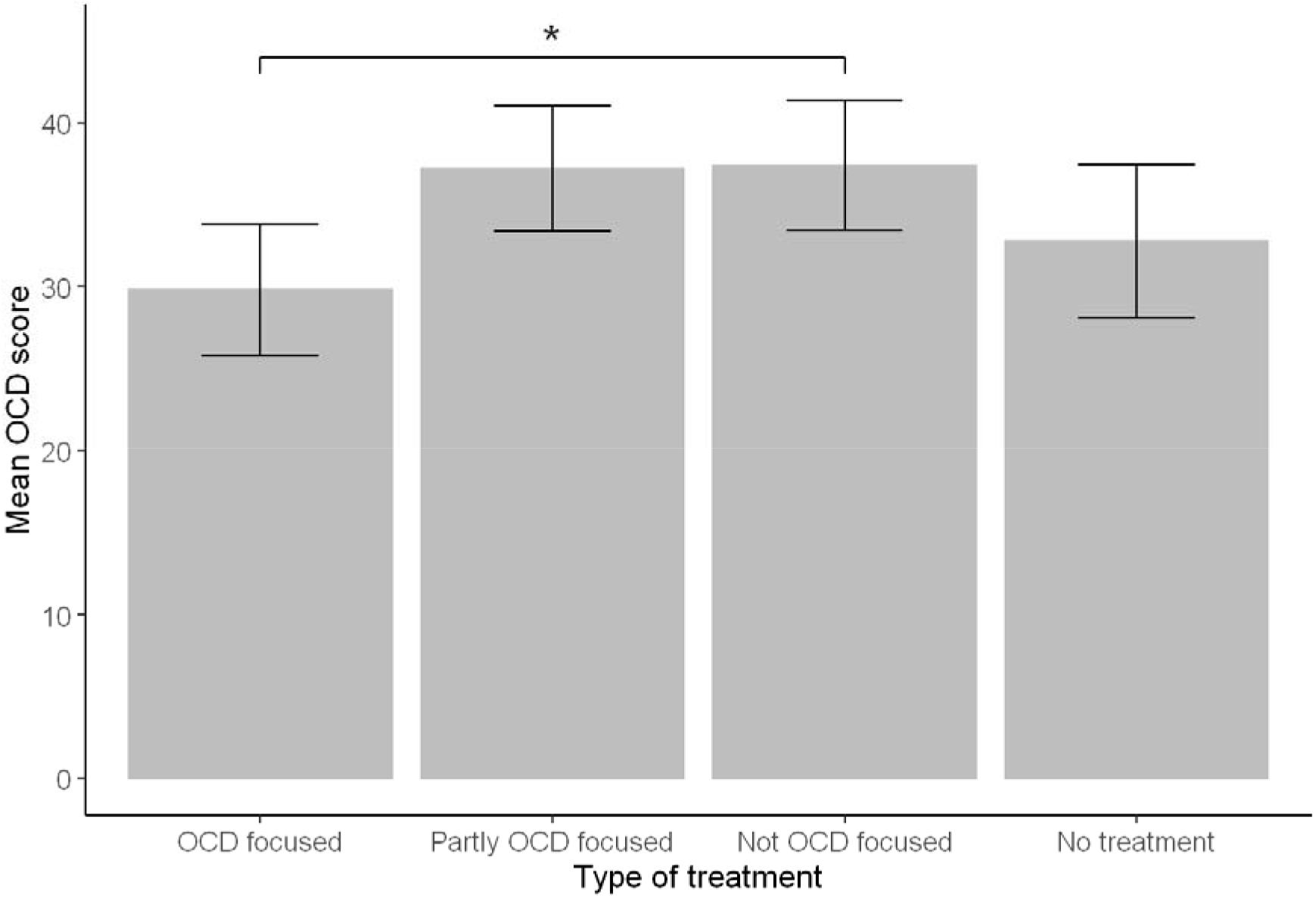
Mean OCD score by type of treatment received. *Note*. OCD focused *n* = 48, Partly OCD focused *n* = 53, Not OCD focused *n* = 50, No treatment *n* = 35. OCD score measured using OCI-R.

Following this, a mixed-model ANCOVA was used to examine whether scores on different OCD subscales varied according to whether the treatment was fully focused on OCD (*n* = 48) or only partly focused on OCD (*n* = 53). Anxiety and depression were included as co-variates. There was a main effect of subscale on OCD score (*F*(5, 485) = 3.12, *p* = .009). Hoarding scores were significantly lower than all other subscale scores apart from neutralising (mean differences - 4.19 to – 1.34, *p*.s < .01), obsessing scores were significantly higher than all other subscales (mean differences 4.19 to 2.23, *p*.s < .001), and checking scores were significantly higher than neutralising scores (mean difference = 1.17, *p* = .02). There were no other differences between subscale scores. There was also a main effect of treatment type (*F*(1, 97) = 6.65, *p* = .01), such that OCD treatment (*M* = 5.00) was associated with significantly lower subscale scores than treatment that was only partly focused on OCD (*M* = 6.22). There was a significant interaction between subscale and treatment type (Figure 3; *F*(5, 485) = 3.66, *p* = .003). In order to examine this interaction in more detail, six univariate ANCOVAs with a Bonferroni-corrected alpha level of .008 were conducted to examine the effect of treatment type on each subscale after controlling for anxiety and depression. It was found that OCD-focused treatment was significantly better than part OCD-focused treatment on hoarding (*F*(1, 97) = 12.77, *p* = .001), neutralising (*F*(1, 97) = 9.48, *p* = .003), and ordering (*F*(1,97) = 7.64, *p* = .007). No significant effect of treatment type was found for checking, obsessing or washing (all *p*.s > .05).

**Figure 3.**
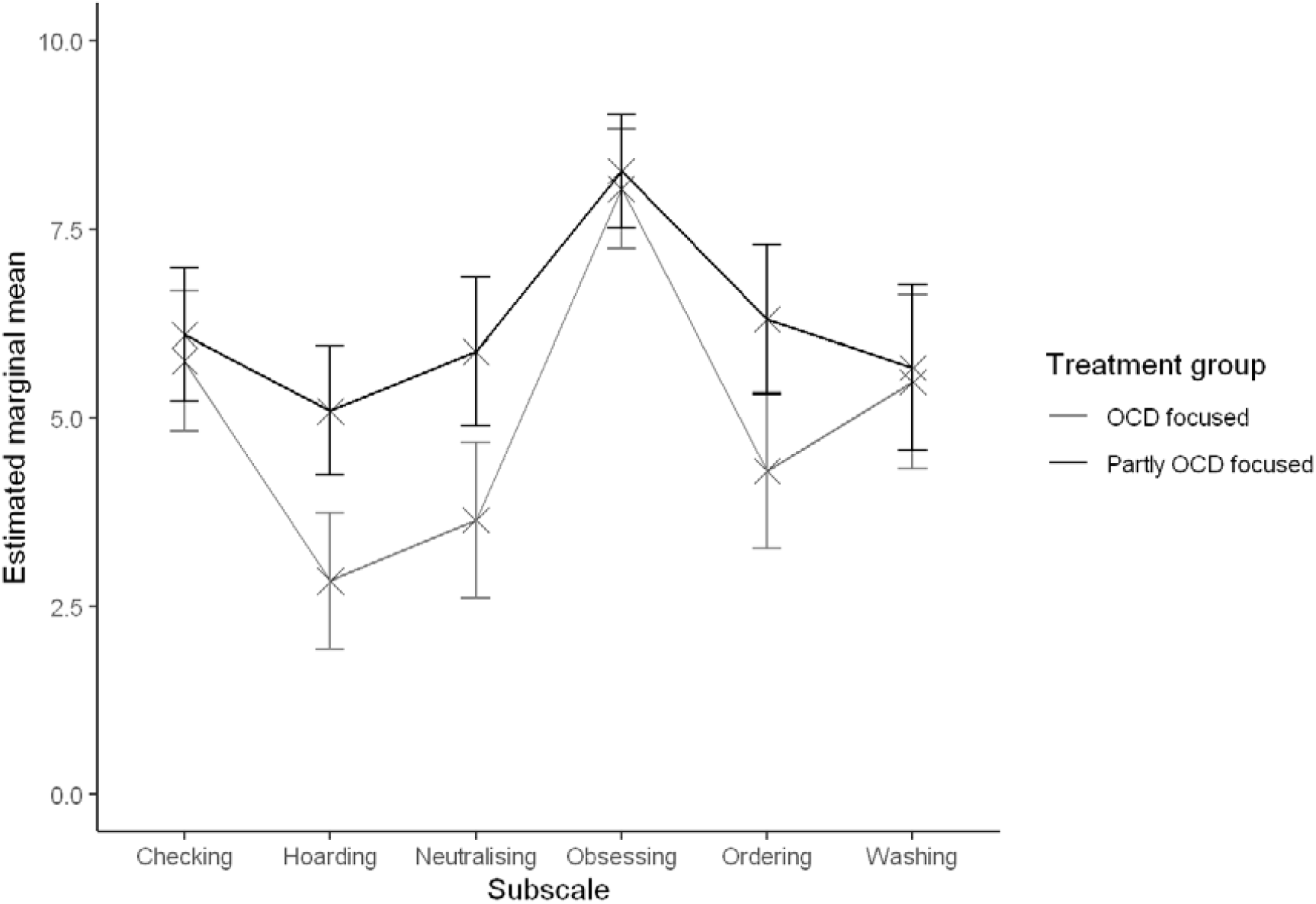
Estimated marginal mean scores on OCD subscales by treatment group. *Note*. Covariates appearing in the model are evaluated at anxiety = 11.04, depression = 7.71. Error bars represent 95% confidence intervals. OCD-focused treatment *n* = 48, part OCD-focused treatment *n* = 53.

### Self-reported treatment response

Participants who had received treatment at least partly focused on OCD were asked to indicate how their current symptoms compared to their symptoms before they had any treatment (Figure 4). This was measured on a scale from 0 to 100, with 0 indicating that symptoms were worse before treatment than now, 50 indicating that the symptoms are about the same before treatment as now, and 100 indicating that the symptoms are worse now than they were before treatment. The median score was 15.5 (IQR: 6.0 – 40.0), indicating that most participants felt that their OCD symptoms were better at the time of the survey than they had been before they first received treatment. It was found that less improvement on this treatment response measure was correlated with more medications tried *r*_*S*_(63) = .39, *p* = .001. Multiple linear regression was then used to examine whether the measure of treatment response was associated with any of the six OCD subscales, or comorbid anxiety or depression. The full model explained 13.2% of the variance in treatment response, though it did not reach significance (*F*(8,101) = 1.76, *p* = .09). Higher scores on the neutralising subscale were associated with less self-perceived improvement in symptoms (*B* = 1.84, 95% CI = 0.16, 3.52, *p* = .03). None of the other predictor variables were significantly associated with treatment response.

**Figure 4.**
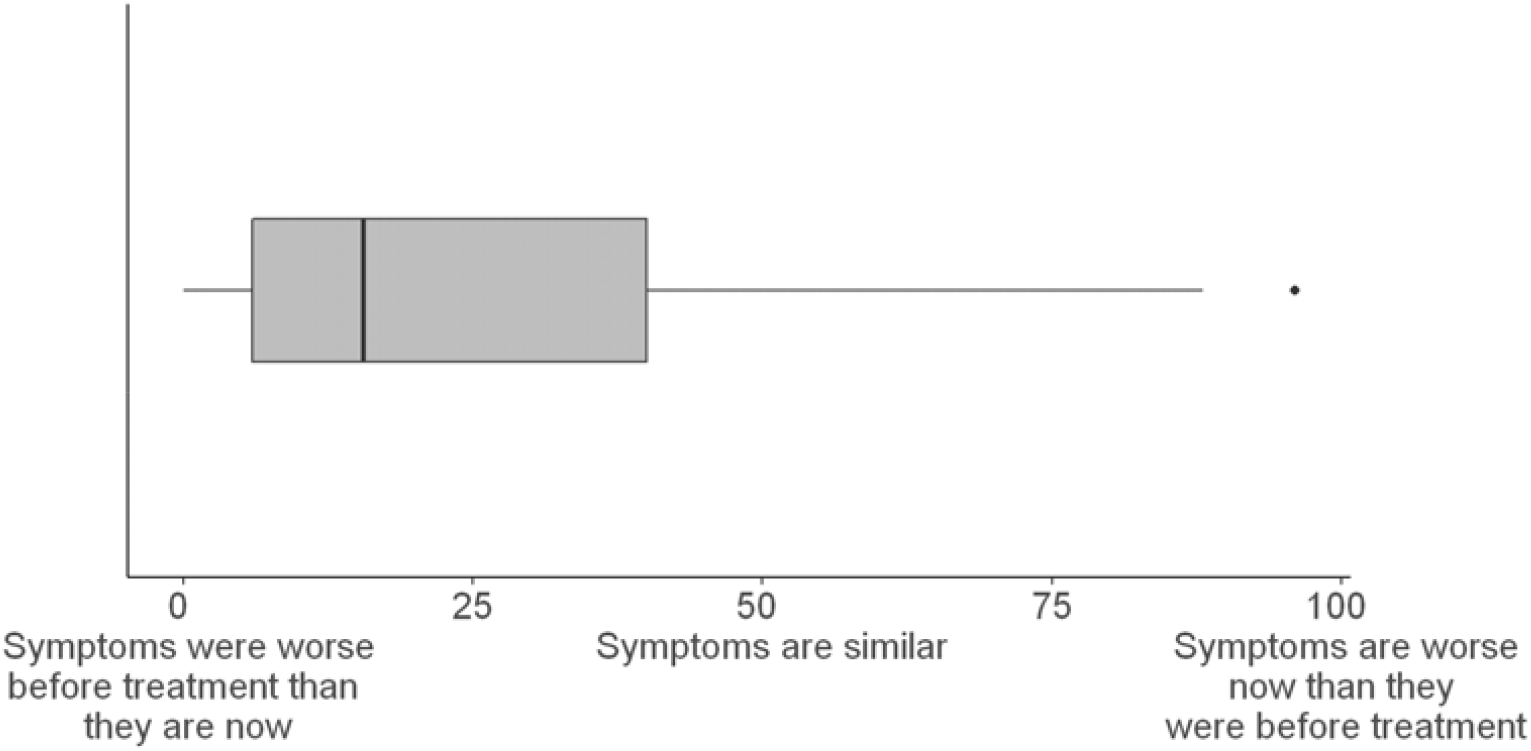
Self-reported symptom change from pre-treatment to present day. *Note. n* = 108.

### Treatment received by people diagnosed with OCD

Amongst participants with a diagnosis of OCD, a majority (54.4%) had received treatment focused on OCD. A further 33.3% had received mental health treatment which was partly focused on OCD, and 7.4% had received mental health treatment which was not focused on OCD at all. A small percentage (4.4%) had not received any mental health treatment.

Diagnosed participants who had received treatment that was at least partly focused on OCD were asked whether they had received cognitive-behavioural therapy/exposure and response prevention therapy for OCD, and whether they had received medication for OCD. Table 1 indicates the number of participants per treatment combination, along with their OCD scores at the time of the survey. Amongst those who had received medication for OCD (*n* = 56), the mean number of medications tried was 2.5 (*SD* = 1.6), the median was 2 and the maximum was 8.

**Table 1.** OCD measures of diagnosed participants by experience of psychotherapy and/or medication

| **Treatment group** | **n** | **Percentage in treatment group** | **Mean (SD) OCI-R** | **Median OCI-R** | **Percentage clinical score** |
| --- | --- | --- | --- | --- | --- |
| Both CBT/ERP and medication | 45 | 58.2% | 31.6 (15.5) | 29 | 73.3% |
| CBT/ERP without medication | 16 | 21.5% | 28.7 (12.0) | 26.5 | 75.0% |
| Medication without CBT/ERP | 10 | 12.7% | 35.8 (13.2) | 36 | 90.0% |
| Neither | 6 | 7.6% | 40.3 (15.0) | 44 | 83.3% |

## Discussion

The present research sought to document the treatment experiences of a sample of people with OCD. Most people reported OCD symptoms which fluctuate in severity over time. Irrespective of what treatment they had previously received, the vast majority of participants scored above the clinical cut-off for OCD at the time of the survey. Notably, this included 73.3% of those who had been diagnosed with OCD and been given both CBT/ERP and medication to treat it. Despite this, most participants felt that their OCD symptoms were less severe now than they had been prior to starting any treatment.

Treatment focused on OCD was associated with fewer OCD symptoms when compared to general (non-OCD) mental health treatment. Treatment focused on OCD was also associated with lower symptoms of hoarding, ordering and neutralising when compared to treatment that was only partly focused on OCD. Symptoms of checking, washing and obsessing did not vary according to treatment type. A tentative interpretation of this may be that practitioners who do not specialise in OCD (i.e. those that deliver treatment partly focused on OCD) may be better able to treat the more widely-known symptoms of OCD, such as checking and washing, than less well-known symptoms, such as neutralising (Pedley et al., 2019). Either way, this set of findings indicates the need for specialist treatment of OCD that goes beyond general mental health treatment (Sookman et al., 2021).

These findings should be considered in light of the important caveat that everyone in the present sample responded “yes” when asked if they considered themselves to have OCD. Individuals whose OCD has been successfully treated are therefore likely to be absent from these data, as those who responded “no” were not asked questions about OCD treatment. Future work should thus ask whether participants had OCD in the past. In addition, as the data are cross-sectional, it is not possible to measure the extent to which OCD symptoms changed from pre-to post-treatment. Consequently, although the current OCD scores were similar across treatment groups, it is possible that pre-treatment scores were higher in one group, and that current parity of symptom severity with the other treatment groups actually reflects greater symptom improvement from baseline. This explanation could also account for the finding that most participants felt that their symptoms had improved after treatment.

Nevertheless, the present findings add to the existing evidence of unmet need in OCD care (Kirkham et al., 2022; Sookman et al., 2021; Vuong et al., 2016), as most of those who had received treatment still experienced clinical levels of OCD. Practitioners should be mindful of the chronic nature of OCD, and the likelihood that one course of CBT/ERP may be insufficient over the long term (Visser et al., 2014). Similarly, researchers should focus more on the factors involved in treatment resistance and relapse, in order to develop more effective treatments that respond not only to randomised control trials but also to people’s lived experience over time (Pittenger & Bloch, 2014).

## Data Availability

De-identified data produced in the present study are available upon reasonable request to the authors.

